# An exploratory study on the correlation of population SARS-CoV-2 cycle threshold values to local disease dynamics

**DOI:** 10.1101/2021.02.16.21251844

**Authors:** Chak Foon Tso, Anurag Garikipati, Abigail Green-Saxena, Qingqing Mao, Ritankar Das

**Author notes:** Corresponding author: **Email:**, 12333 Sowden Rd., Ste B, PMB 65148, Houston, Texas 77080-2059, (510) 826 - 9508.

## Abstract

**Introduction:** Despite limitations on the use of cycle threshold (CT) values for individual patient care, population distributions of CT values may be useful indicators of local outbreaks.

**Methods:** Specimens from the greater El Paso area were processed in the Dascena COVID-19 Laboratory. Daily median CT value, daily transmission rate R(t), daily count of COVID-19 hospitalizations, daily change in percent positivity, and rolling averages of these features were plotted over time. Two-way scatterplots and linear regression evaluated possible associations between daily median CT and outbreak measures. Cross-correlation plots determined whether a time delay existed between changes in the daily median CT value and measure of community disease dynamics.

**Results:** Daily median CT was negatively correlated with the daily R(t), the daily COVID-19 hospitalization count (with a time delay), and the daily change in percent positivity among testing samples. Despite visual trends suggesting time delays in the plots for median CT and outbreak measures, a statistically significant delay was only detected between changes in median CT and COVID-19 hospitalization count.

**Conclusions:** This study adds to the literature by analyzing samples collected from an entire geographical area, and contextualizing the results with other research investigating population CT values.

## INTRODUCTION

As of February 16, 2021, the SARS-CoV-2 virus has infected more than 109 million people around the world, and has been implicated in 2.41 million deaths (1). In the United States (US) alone, more than 486,500 deaths have been attributed to coronavirus disease 2019 (COVID-19) (1). Reverse transcription polymerase chain reaction (RT-PCR) tests have become the predominant method for COVID-19 surveillance and diagnostic testing due to their high sensitivity, high specificity and rapid turn-around time compared to viral culture (2,3). RT-PCR tests detect viral genetic material in biological samples (4). The cycle threshold (CT) value represents the number of PCR cycles required in order to detect a positive signal (5). The CT value is inversely related to the viral load, such that a 3.3 increase in CT value indicates an approximately 10-fold decrease in the amount of viral genetic material present in a sample (5). COVID-19 RT-PCR tests are generally considered positive only if they generate a result with a CT value lower than the recommended cut-off; in the US, the Food and Drug Administration (FDA) has approved emergency use authorizations (EUAs) for tests for which CT values < 37 can be considered positive (6).

CT values are lowest, indicating a larger amount of viral genetic material, early in the disease course. Indeed, numerous studies have reported that CT values tend to be highest prior to or in the earliest days following the onset of symptoms, and decline as disease progresses (7–11). Lower CT values have been directly linked with higher infectivity, as shown by the ability to cultivate live SARS-CoV-2 from samples (7,11–14) and the number of individuals infected by an index case (15). In one study, SARS-CoV-2 could be cultivated from over 70% of samples with a CT value < 25, but from less than 3% of samples with CT value ≥ 35 (16). CT values have also been directly correlated with disease severity and mortality, such that the CT values tend to be lower in patients with more severe disease presentations (13,17–19) and in hospitalized patients who ultimately die from COVID-19 (13,20).

However, there are significant limitations to the use of CT values for prognostication and treatment planning at the level of the individual patient. Critics have noted that there may be significant variability in CT values based on the quantity of biological material collected on a testing swab, as well differences in RT-PCR reagents, equipment and standards between laboratories (21). CT values may also vary based on the gene target selected for RT-PCR, or even on the assay used to detect the same gene target (22). In addition, RT-PCR only detects the presence of viral material, and is unable to distinguish between live virus and viral debris, which may linger for an extended period once an individual is no longer infectious (17). CT values, as a semi-quantitative measure of how much viral nucleic acid is present, are similarly limited. As a result of these constraints, clinicians and researchers continue to debate the utility of CT values for informing healthcare choices for individuals (e.g. (5,21,22)).

Despite the limitations on the use of individual-level CT values, measures of CT values across a population may provide a useful measure of COVID-19 dynamics in the community. As CT values have been correlated with disease stage and infectivity, a higher proportion of low CT values in testing samples from the population may reflect epidemic growth in the community (23). Preliminary analyses of simulation and surveillance testing data suggest that decreases in the distribution of CT values in a population, as measured by the median CT value, may precede a local outbreak, such that the median CT value may be a useful tool in predicting a surge (23,24). The present study describes an exploratory analysis of potential correlations between median CT values and COVID-19 disease dynamics, operationalized as percent positivity, transmission rate and COVID-19 hospitalizations.

## METHODS

### Sample Selection

The samples in this study were collected between September 15th, 2020 and January 11th, 2021 as part of the ongoing diagnostic evaluation services provided by Dascena, Inc to residents in the state of Texas. In the greater El Paso area, a contractor for the El Paso Department of Public Health sends over 90% of collected samples to the Dascena COVID-19 Laboratory in Houston, Texas. The Pearl Independent Institutional Review Board (IRB) approved this study (IRB Protocol 21-DASC-127).

This study included nasopharyngeal swabs, salivary samples, an anterior nares swabs sample, and samples for which the type of biological specimen was not specified. The overwhelming majority of samples were nasopharyngeal swabs. All biological samples were sent to the Clinical Laboratory Improvement Amendments (CLIA)-certified Dascena Laboratory. All samples were analyzed with TaqPath COVID-19 Combi Kit (Thermo Fisher Scientific, Waltham, Massachusetts), with extraction performed with a MagMAX RNA Isolation Kit (Thermo Fisher Scientific, Waltham, Massachusetts). Three gene targets are used by these assays, and may be the source of a positive result: the nucleocapsid (N) gene, the spike (S) gene and the open reading frames (ORF1ab) gene (25). RT-PCR was only run once on any unique sample. For each RT-PCR test, the CT value was recorded. Only samples that produced a valid CT value for a positive COVID-19 test (i.e., at least 2 genes generating a positive signal with a CT value ≤ 37) were used to determine the daily median CT value and in subsequent correlation analyses.

### Data Processing and Measures

The following demographic data were available for testing samples: age, sex, race, ethnicity, and zip code of residence. Testing samples from the greater El Paso area were selected based on the zip codes listed as part of the El Paso metropolitan statistical area (MSA) by the US Department of Labor, Office of Workers Compensation Program (26). Daily percent positivity rate was calculated among all samples tested by Dascena from the greater El Paso area.

The effective reproduction number or transmission rate R(t), was derived using the open-source algorithm from COVID-19 tracking website rt.live (27). The algorithm is a Python script based on a Bayesian Estimation Model developed by Bettencourt & Ribeiro (28), with slight modification to introduce gaussian noise to the prediction. Daily new COVID-19 case data from individual counties were obtained from the COVID-19 Dashboard by the Center for Systems Science and Engineering at Johns Hopkins University (1), grouped by MSA, and fed into rt.live’s algorithm to generate a time series for R(t). The daily number of individuals hospitalized with COVID-19 in the El Paso area was derived from publicly available data produced by the Texas Department of State Health Services that are grouped by trauma service area (29).

### Comparative Analysis of Population-level CT Values

In order to contextualize the results, a focused literature search was performed for peer-reviewed publications and pre-print manuscripts on the use of CT values measurements across a population as a means of predicting or monitoring COVID-19 outbreaks. Three pre-prints were identified (23,24,30). The datasets from the present study and the pre-prints were then compared in terms of: source population; type of testing; sample size; biological sample types include; duration of study period; gene target(s) of RT-PCR tests; CT-based value(s) measured; metrics used to measure COVID-19 outbreaks; and the outcomes of study.

### Statistical Analysis

All analyses were conducted in Python (31) using the following packages: pandas, matplotlib, plotly, scipy and statsmodels. The daily median CT value among Dascena test samples, the daily R(t) in the El Paso MSA, and the daily count of hospitalized individuals with COVID-19 in El Paso were plotted over time. Rolling 7-day averages of the daily median CT value (with a minimum 5 days of data present in the window), the daily R(t), the daily number of COVID-19 hospitalizations, and the daily percent positivity among samples from El Paso sent to the Dascena Laboratory were also plotted over time. To better capture the dynamic change in percent positivity among Dascena test samples, the daily change in percent positivity was calculated from the 7-day rolling average for days with more than 200 total tests performed by the Dascena Laboratory. If fewer than 200 tests were performed on a particular day (e.g., due to holiday shut down of collection sites), the percent positivity from the previous day was carried forward. The daily change in percent positivity was then plotted over time.

Scatterplots and linear regression were used to evaluate possible associations between the daily median CT value (N gene) and daily R(t), between the daily median CT value (N gene) and the daily count of COVID-19 hospitalizations, and between the daily median CT value (N gene) and the daily change in percent positivity among samples processed by Dascena. Since a significant time delay was observed between changes in the daily median CT value (N gene) and the daily count of COVID-19 hospitalizations, a time lag of 33 days was applied to the hospitalization data prior to creating the scatterplot and conducting linear regression. The median CT value based on the N gene was selected because it has previously been cited in research on population CT values (24,30).

In order to evaluate whether a time delay existed between changes in the daily median CT value (N gene) and community outbreaks, cross correlation plots were constructed between daily median CT value and daily R(t), between daily median CT value and the daily count of patients hospitalized with COVID-19, and between daily median CT value and the daily change in percent positivity. In brief, a cross-correlation coefficient was obtained by dividing the correlation between two signals to the product of auto-correlation of each of the two signals. The argmax of the cross-correlation coefficient is the dominant lag time between the two signals. As the purpose of the present analysis was to investigate how the trough of daily median CT correlated with the peak of the other signals, to aid visualization the following modifications were made: (1) for each signal, the z-score was used instead of the absolute value; (2) the negative value of the z-score of daily median CT value was used to ensure a positive peak in the cross correlation plots; (3) 20% of positive samples were randomly sampled five times each day to estimate the variation in the cross-correlation between daily median CT value and epidemiological signals. A 1-sample t-test was used to determine if the mean lag differed statistically significantly from the zero.

Pairwise comparisons were performed with Pearson’s correlation (significance P < 0.05) to determine if any demographic factors associated with testing samples were significantly associated with R(t), COVID-19 hospitalization count, or percent positivity. The following demographic factors were investigated: daily number of tests; daily median age; daily percent samples from men; daily percent samples from individuals indicating White race; daily percent sample from individuals indicating Hispanic ethnicity.

## RESULTS

In the greater El Paso area, 148,410 COVID-19 tests were sent to the Dascena Laboratory for processing, and 36,306 tests were positive. 147,720 (99.54%) of samples were nasopharyngeal swabs, 28 (0.02%) were salivary samples, 1 sample (0%) was an anterior nares swab, and 661 samples were biological specimens (0.45%) for which the type of specimen was not recorded. The median CT value (N gene) for nasopharyngeal samples was 23.14, which differed significantly from the median CT value (N gene) observed for all other sample types of 25.58 (p < 0.05, Mood’s median test). The demographic characteristics of the entire population tested for COVID-19 are presented in **Table 1**.

**Table 1.** Demographic characteristics of population from greater El Paso area with COVID-10 tests submitted to the Dascena Laboratory

|  |  |
| --- | --- |
| Age (Mean, SD) | 36.92 (18.53) |
| **Gender** |  |
| Female | 81,520 (54.93%) |
| Male | 66,270 (44.65%) |
| **Race** |  |
| White | 87,664 (59.07%) |
| American Indian or Alaska Native | 3,744 (2.52%) |
| Black/African American | 3,017 (2.03%) |
| Asian or Pacific Islander | 1,245 (0.84%) |
| Unknown | 52,203 (35.16%) |
| **Ethnicity** |  |
| Hispanic | 127,722 (86.06%) |
| non-Hispanic | 8,632 (5.82%) |
| Unknown | 11,963 (8.06%) |

Variability over time was observed in the median CT values and measures of COVID-19 disease dynamics in El Paso (**Figure 1**). As predicted based on the *a priori* hypothesis, the daily median CT was negatively correlated with the daily R(t), daily count of COVID-19 hospitalization (with a time delay), and the daily change in percent positivity among testing samples in the greater El Paso area (**Figure 2**).

**Figure 1.**
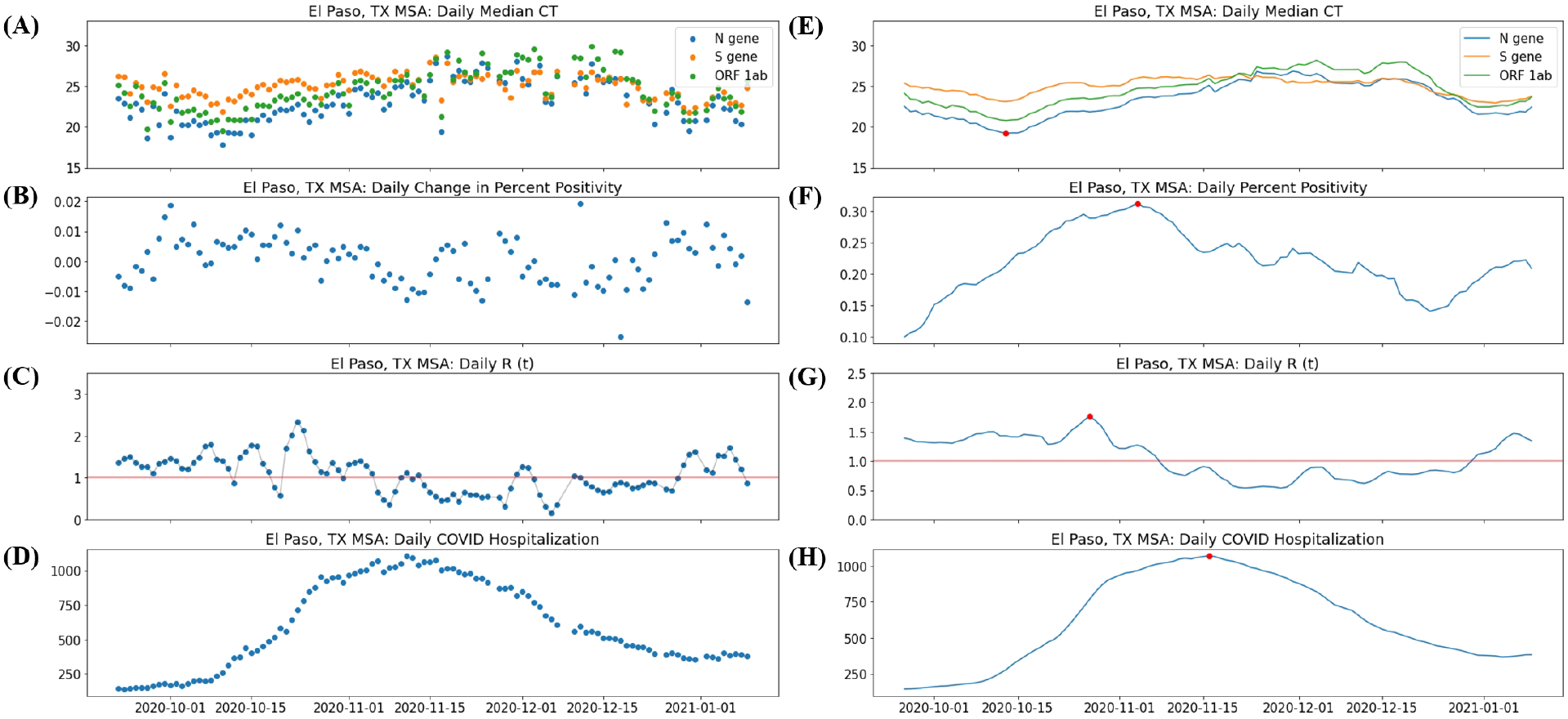
**(A)** Daily median cycle threshold (CT) value, **(B)** daily change in percent positivity, **(C)** daily transmission rate, R(t), **(D)** daily count of individuals hospitalized with COVID-19; **(E)** 7-day rolling average of the daily median CT value, **(F)** 7-day rolling average of percent positivity, **(G)** 7-day rolling average of daily R(t), and **(H)** 7-day rolling average of individuals hospitalized with COVID-19, in the greater El Paso area. Red line in **(C)** and **(G)** signifies R (t) = 1.Red dots are global minimum for smoothed CT value; and global maxima for smoothed epidemiological indicators.

**Figure 2.**
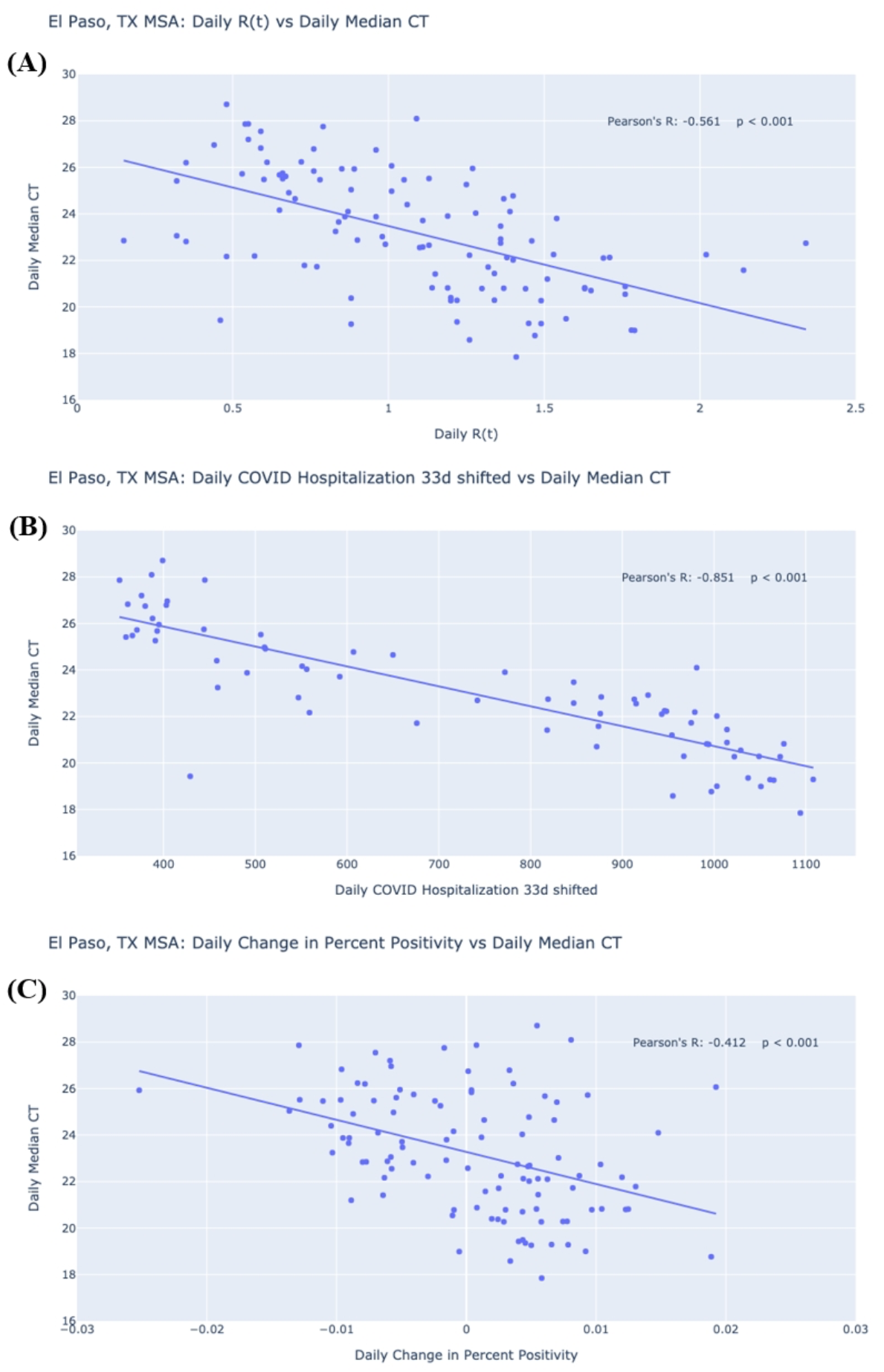
Linear regression and scatter plot of **(A)** daily median cycle threshold (CT) values versus daily transmission rate R(t), **(B)** daily number of individuals hospitalized with COVID-19, and **(C)** daily change in percent positivity for greater El Paso area

A 32-34 day shift was observed between median CT and the daily count of individuals hospitalized with COVID-19 (**Figure 3**). While visual inspection of the daily median CT, daily R(t) and percent positivity plots over time (**Figure 1**) suggested that peaks in R(t) and percent positivity followed a trough in median CT, no statistically significant time delays were detected between median CT and change in percent positivity or R(t).

**Figure 3.**
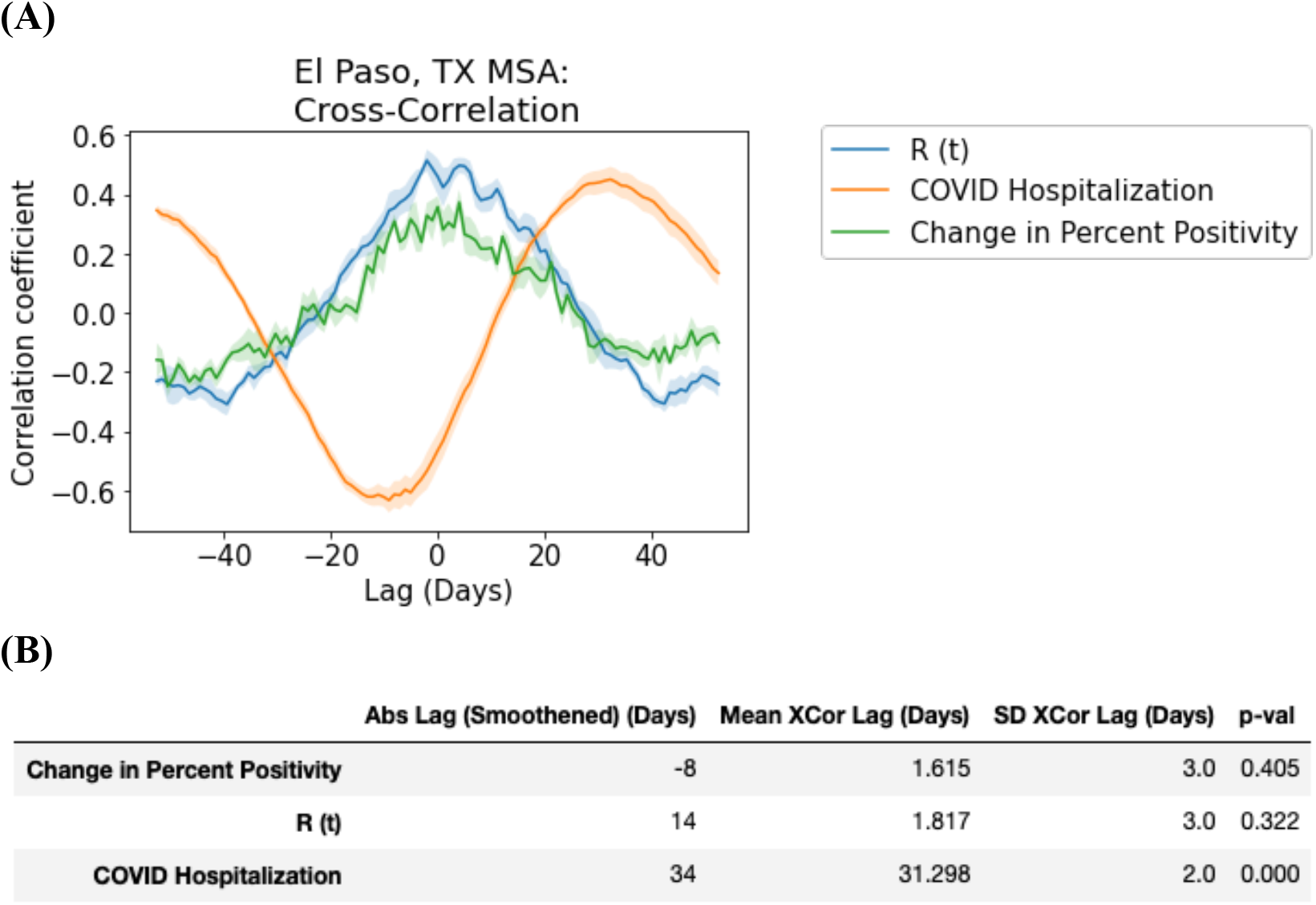
**(A)** Cross-correlation plot of daily median cycle threshold (CT) values versus daily transmission rate R(t), daily number of individuals hospitalized with COVID-19, and daily change in percent positivity for the greater El Paso area. Solid lines represent the mean, while shaded areas indicate standard deviation across 5-fold sampling. **(B)** Lag between epidemiological signals and daily median CT (N gene). Abs. Lag (smoothed) is the absolute time difference between the peak of each epidemiological signal and the trough of daily median CT with a 7-day rolling average (red dots of figure 1b). Mean XCor Lag and SD XCor Lag represent the mean and standard deviation, respectively, among lags determined by the 5-fold sampling of daily median CT and cross-correlation. P-val shows the p-value of whether the cross-correlation between daily median CT and each of the epidemiological signals is statistically different from zero by 1-sample t-test.

Pairwise comparisons revealed that some demographic factors of testing samples were associated with COVID-19 outbreak measures (**Table 2**). No other factors, including the number of tests performed, median age, or percentage of tests from male, Hispanic or White individuals each day, were significantly correlated with daily R(t), daily difference in positivity rate or daily count of COVID-19 hospitalizations in the El Paso area.

**Table 2.** Variables correlated with measures of COVID-19 outbreak

| Measure of COVID-19 outbreak | Correlated Variable | Correlation Coefficient | Significance |
| --- | --- | --- | --- |
| Daily R (t) | Daily Median Age | -0.332 | $p < 0.001$ |
| Daily R (t) | Daily percentage of samples from White individuals | 0.3866 | $p < 0.001$ |
| Daily Change in Percent Positivity | Daily percentage of samples from White individuals | 0.251 | $p < 0.01$ |
| Daily Change in Percent Positivity | Daily percentage of samples from Hispanic individuals | 0.265 | $P < 0.01$ |
| Daily COVID Hospitalization (33d shifted) | Daily Median Age | -0.451 | $P < 0.001$ |
| Daily COVID Hospitalization (33d shifted) | Daily percentage of samples from White individuals | 0.567 | $P < 0.001$ |

The dataset in this study was substantially larger than those used in comparator studies, but differed in that it was not a surveillance sample. Instead, this study used samples from individuals who required testing due to the presence of COVID-19 symptoms, or who required testing in the absence of symptoms (e.g. for work or travel clearance). Median CT value was the most common measure of population distribution of CT values across research studies to date, and R(t) and percent positivity were the most common outbreak measures (**Table 3**).

**Table 3.**
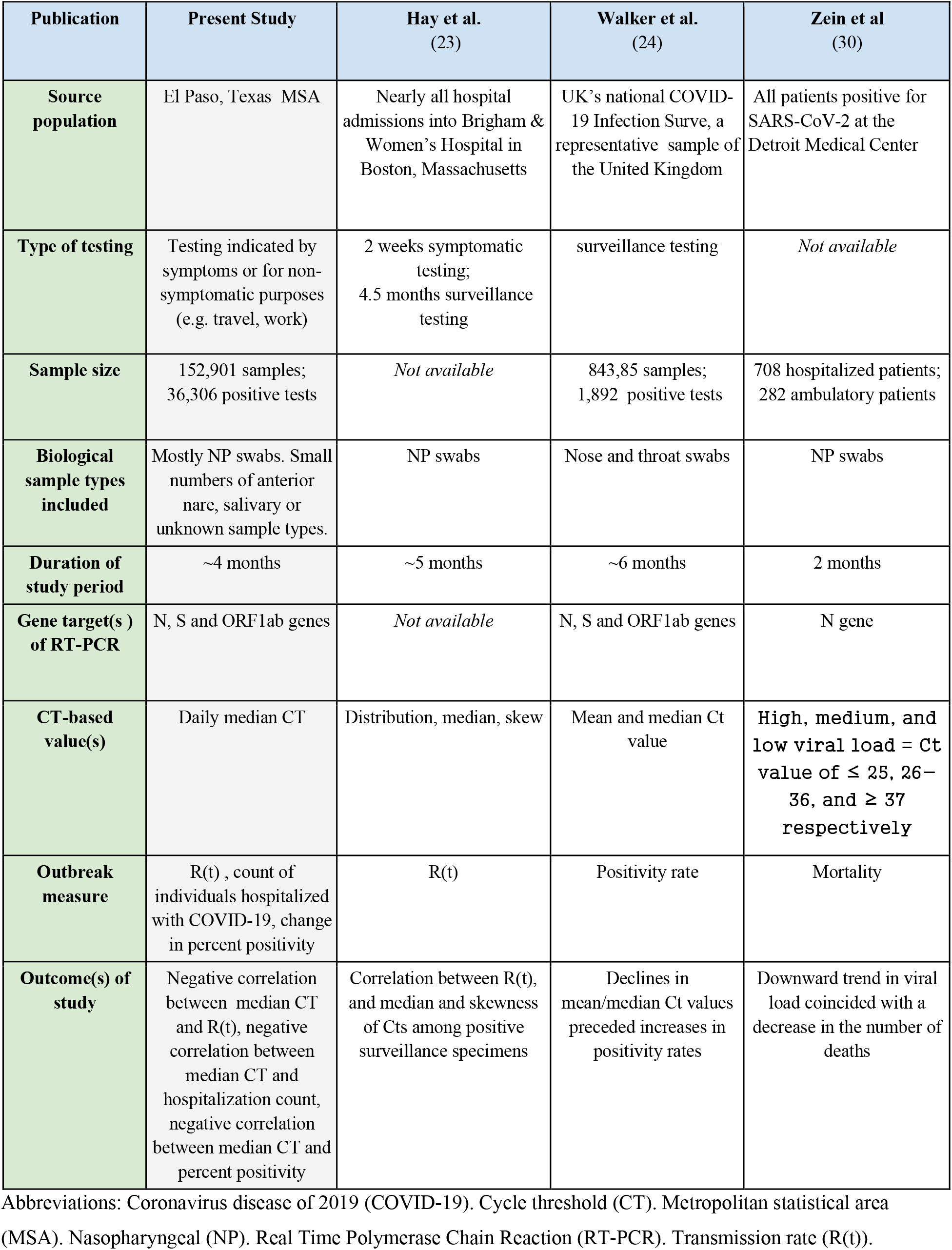
Comparison of studies that examine Cycle Threshold values at the population level.

## DISCUSSION

At present, surges are largely predicted based on observed local case and mortality rates, which may lag by several weeks behind changes in transmission rates or be obscured by changes in testing capacity (23). Given the ubiquitous availability of CT data and the pressing nature of the pandemic, interest has risen in exploring the possibility that the population distributions of CT values can be used as indicators for local outbreaks. The current study adds to the growing literature on this topic by providing an analysis of median CT values from samples collected from an entire geographical area, and contextualizing the results with a comparison to other research investigating the application of population CT values.

In the greater El Paso area, daily median CT values were found to be negatively correlated with the daily percent positivity among samples, the daily R(t) extracted from community case rates and the daily count of COVID-19 hospitalizations (with a delay). Of note, these associations were not observed in supplementary analyses (**Supplementary Figure 1; Supplementary Figure 2; Supplementary Figure 3**) conducted for different Texas MSAs for which substantially fewer tests, covering a smaller proportion of the population (**Supplementary Table 1**), were processed. There appeared to be greater day-to-day variability in the median CT values over time rather than consistent trends in the MSAs evaluated in supplementary analyses, potentially reflecting differences in the strength of the signal that could be detected. In addition, substantial differences in the study populations may have contributed to variable significance of the relationship between median CT value and outbreak measures between study sites. This hypothesis is supported by the observation of significant demographic differences between El Paso MSA and the Texas MSAs evaluated in the supplementary analyses (**Supplementary Table 2**). This observation indicates that certain qualities of datasets used to measure population CT values may be important to their utility to approximate local COVID-19 surges.

Changes in the population distribution of CT values significantly preceded a rise in COVID-19 hospitalizations in El Paso. However, contrary to the *a priori hypothesis* that changes in CT values would precede surges, the cross-correlation plots of median CT value, percent positivity and R(t) did not strongly demonstrate such a relationship. It therefore remains unclear from the data whether changes in the population distribution of CT preceded changes in community transmission, or vice versa. Other studies evaluating population CT values in surveillance samples have reported that changes in CT values may precede traditional signs of an outbreak (23,24). The inclusion of symptomatically indicated tests in the sample population may have influenced this association, such that a decline in CT values may be more closely linked to current case rates.

### Strengths

Strengths of this study include that all RT-PCR analyses were conducted at a single laboratory using standardized testing protocols, and that large samples of positive COVID-19 tests were acquired for the study site. The vast majority (>99%) of samples were nasopharyngeal swabs, such that differences in median CT values based on sample type likely did not impact results. This study was not limited to a single medical center, but included samples collected from an entire geographical area. This research compared median CT values to R(t) and hospitalization count, traditional public health used benchmarks to define surges, providing greater validity than would be possible with only an internal comparison of different metrics of testing sample data. In addition, this study provided a novel examination of the features of RT-PCR testing data which may contribute to and affect the usability of population-level metrics of CT values for predicting disease dynamics in the community.

### Limitations

While the study sample was large, other variables and forms of bias (e.g., sampling bias), may have influenced the results. Indeed, differences in the comprehensiveness of the El Paso dataset versus the supplementary site datasets-or in, other words, the relative proportion of tests conducted by the Dascena laboratory versus other testing providers-may have contributed to skew in the supplementary samples. Future directions for research on population CT values may therefore include analyzing whether significant differences in results can be detected in different sub-samples of tested populations, and evaluating methods to collate CT data across testing providers in a given geographic area.

No data on symptomatology was associated with samples at the time of collection, such that these data do not enable a distinction between samples collected as part of clinical evaluation of symptoms consistent with COVID-19, or for other reasons (e.g., clearance for work or travel). Prior research assessing population distribution of CT values in relation to community outbreaks has explicitly used surveillance samples (23,24). The variability in the observed correlations between median CT and outbreak measures in El Paso versus other testing locations may reflect, in part, variability in the proportion of symptomatically indicated versus non-symptomatically indicated tests in a given location. However, other differences between the testing site populations may also have contributed to the observed variability in the relationship between median CT value and outbreak measures, such as differences in the demographics of the tested population. The research question of whether median CT values derived from all testing data, versus only surveillance testing data, may be reliably used to predict disease outbreaks remains unresolved, and can only be addressed using datasets in which symptomatology at the time of testing or reason for testing may be linked to test results.

The samples used in this study were not collected expressly for the purposes of public health surveillance or research, and so the demographic composition of the sampled population varied day to day. As indicated by **Table 2**, some aspects of the daily demographic composition of the tested population were found to correlate with epidemiological outcomes. Daily variability in the sampled population may therefore translate to variability in the strength of the associations between median CT and measures of disease dynamics. However, these associations may also reflect underlying epidemiological trends, such as the disproportionately high rates of COVID-19 infection among Hispanic individuals (32), including during outbreaks. Additional research with real-world samples may build on this work by further exploring the relevance of demographic factors to the accuracy and utility of population CT measures.

## CONCLUSIONS

As national, state and local authorities continue to refine public health programs to track and contain the spread of SARS-CoV-2, it is imperative to optimize methods for predicting surges in community transmission. Greater lookahead time would enable local and state officials to enact public health policies to mitigate an anticipated surge, and would provide health systems with the opportunity to initiate changes to their standard operating procedures, including activating reserve clinical personnel, procuring additional resources to the extent possible, and converting facilities to support additional patient flow. The population distribution of CT values, as measured by the median CT value, is a potential indicator for local outbreaks, which merits further investigation.

## Data Availability

The dataset analyzed in this study is proprietary and not available for distribution. Additional data pertaining to the analyses performed may be available on reasonable request.

## Author roles

CFT processed the data, adapted the software code and conducted statistical analyses, generated figures, contributed to drafting the manuscript, and participated in critically reviewing and editing the manuscript. AG obtained and organized the data for study, reviewed software and statistical analyses, and contributed to the primary drafting and editing of the manuscript. AGS contributed to critical review of study design and analyses, drafting the manuscript and editing the manuscript. QM and RD formulated the idea for this study, supervised analyses and critically reviewed and edited the manuscript.

## Supplement

**Supplementary Figure 1.**
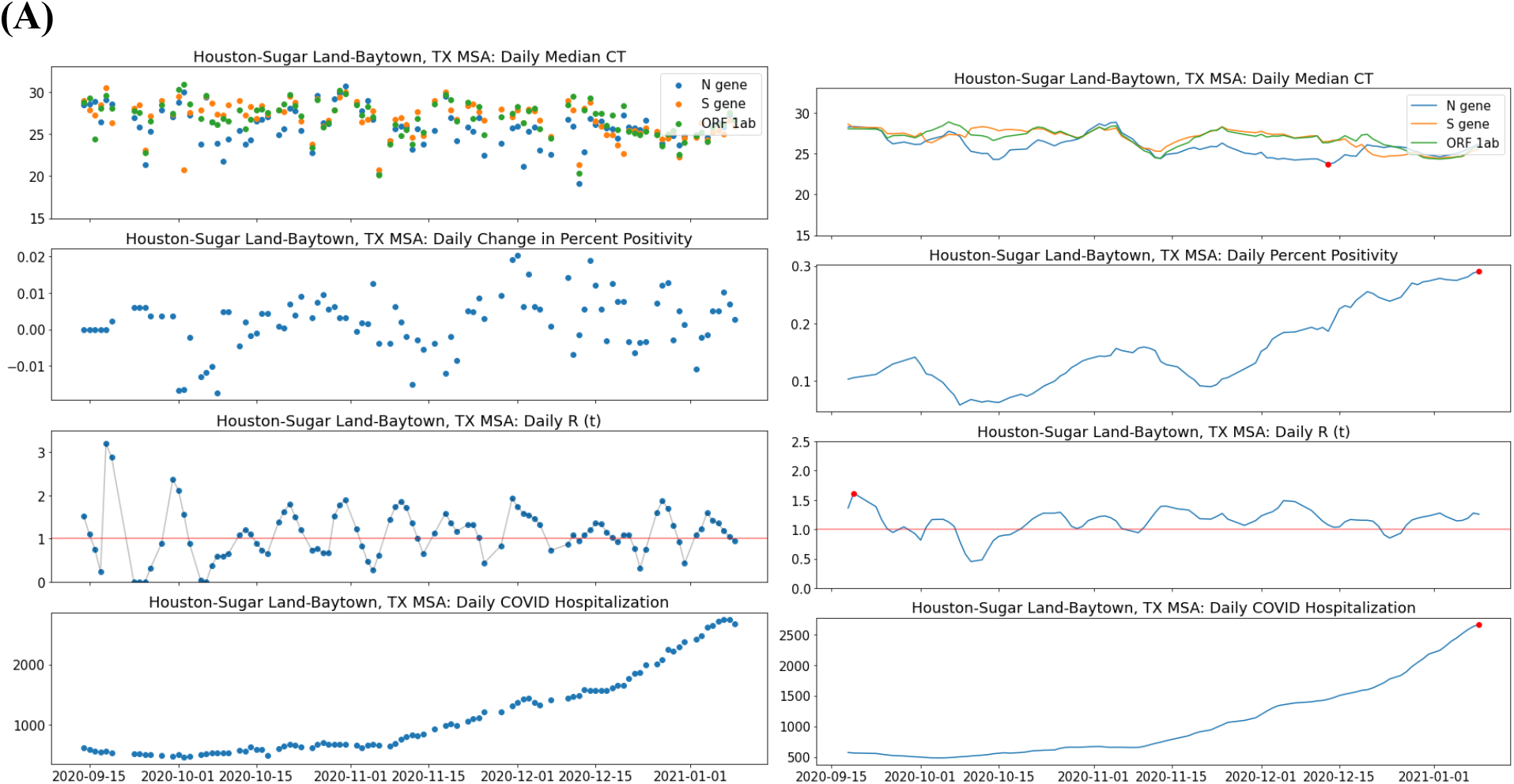
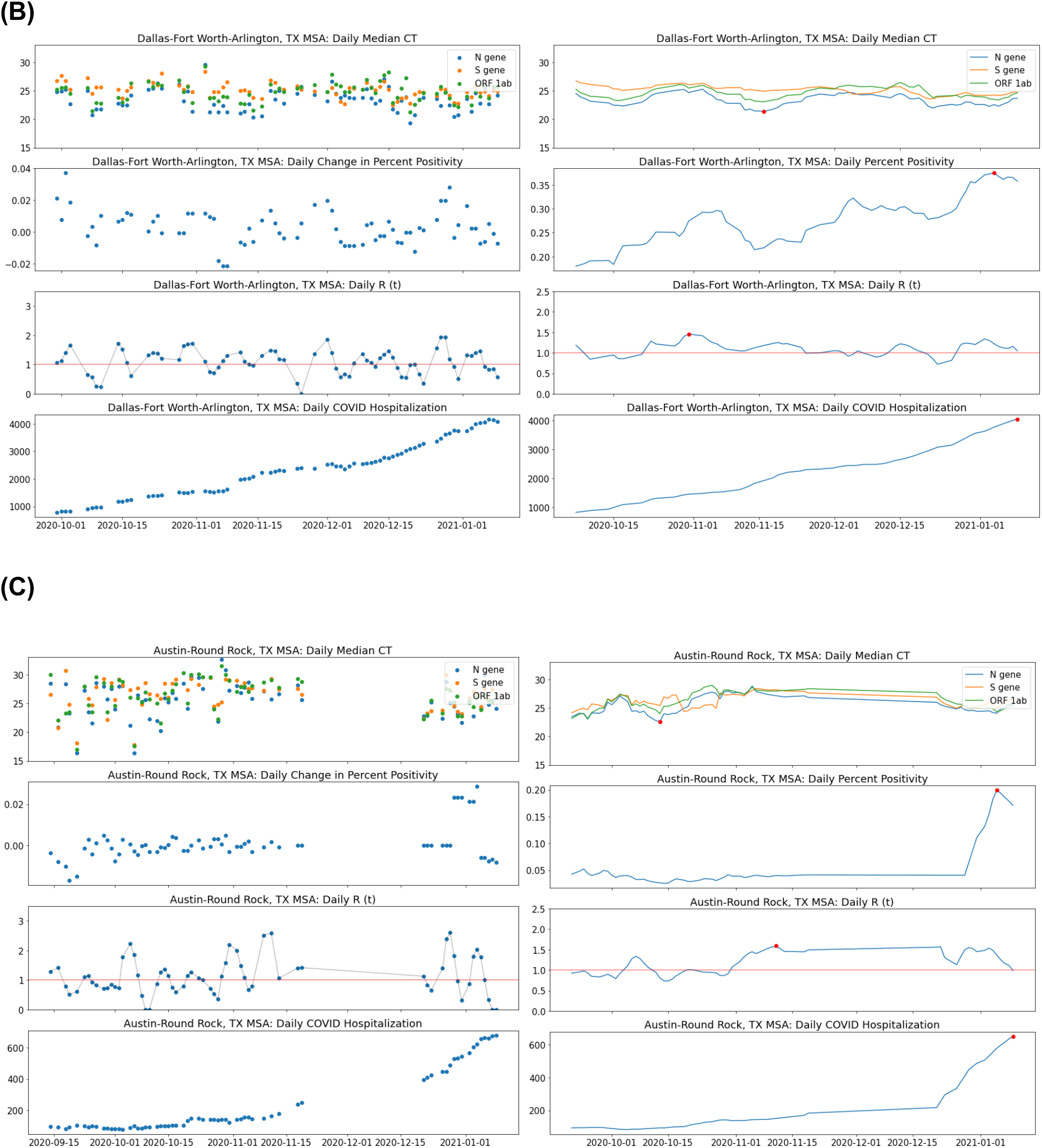
Daily median cycle threshold (CT) value, daily transmission rate R(t), daily number of individuals hospitalized with COVID-19, daily change in percent positivity, 7-day rolling average of the daily median CT value, 7-day rolling average of daily R(t), 7-day rolling average of the number of individuals hospitalized with COVID-19 and rolling average of the percent positivity among Dascena samples in (A) Houston-Sugarland-Baytown Metropolitan Statistical Area (MSA), (B) Dallas-Fort Worth-Arlington MSA, and (C) Austin-Round Rock MSA

**Supplementary Figure 2.**
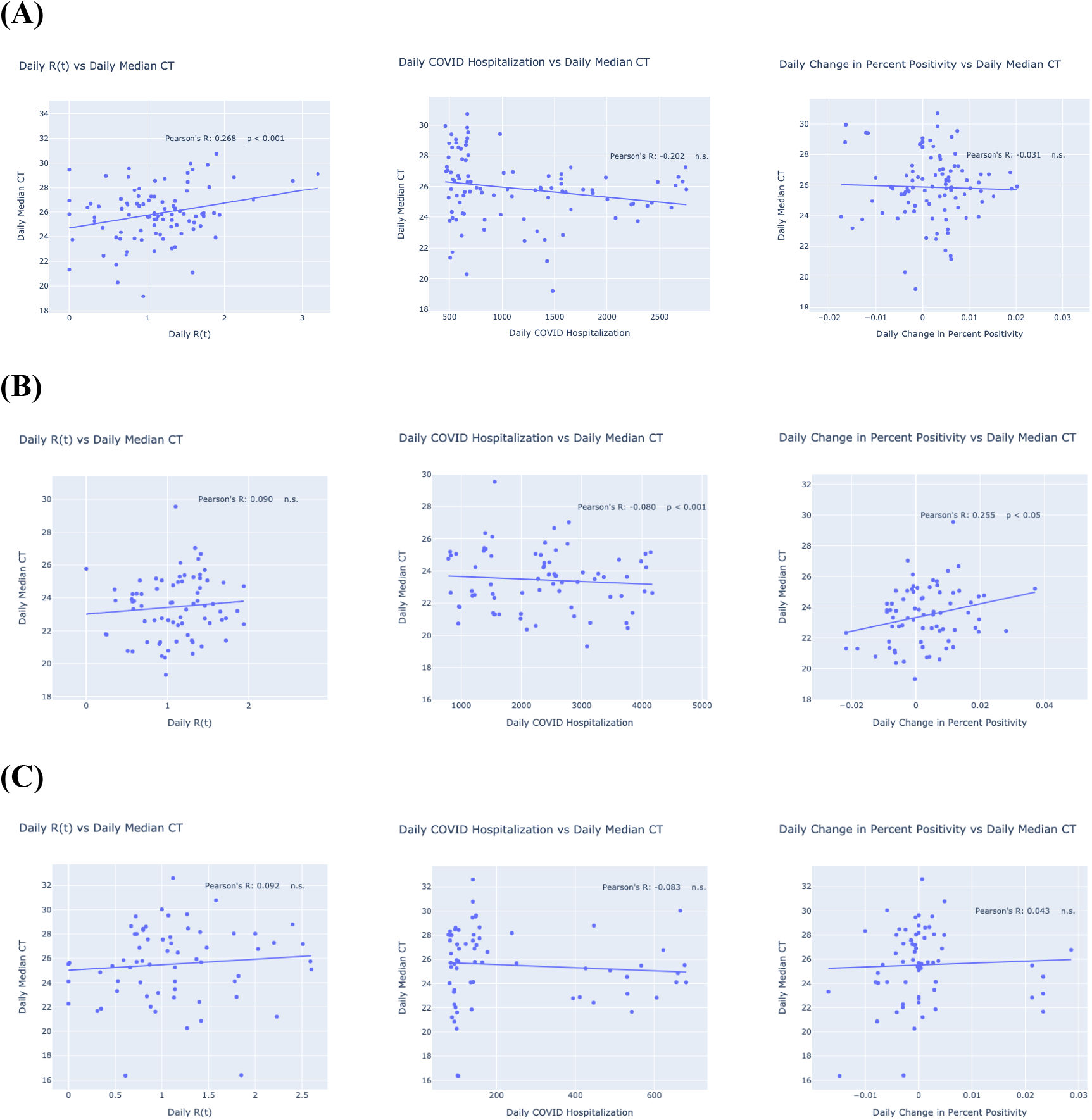
Linear regression and scatterplots of daily median cycle threshold (CT) values versus daily transmission rate R(t), daily number of individuals hospitalized with COVID-19, and daily change in percent positivity for (A) Houston-Sugarland-Baytown Metropolitan Statistical Area (MSA), (B) Dallas-Fort Worth-Arlington MSA, and (C) Austin-Round Rock MSA

**Supplementary Figure 3.**
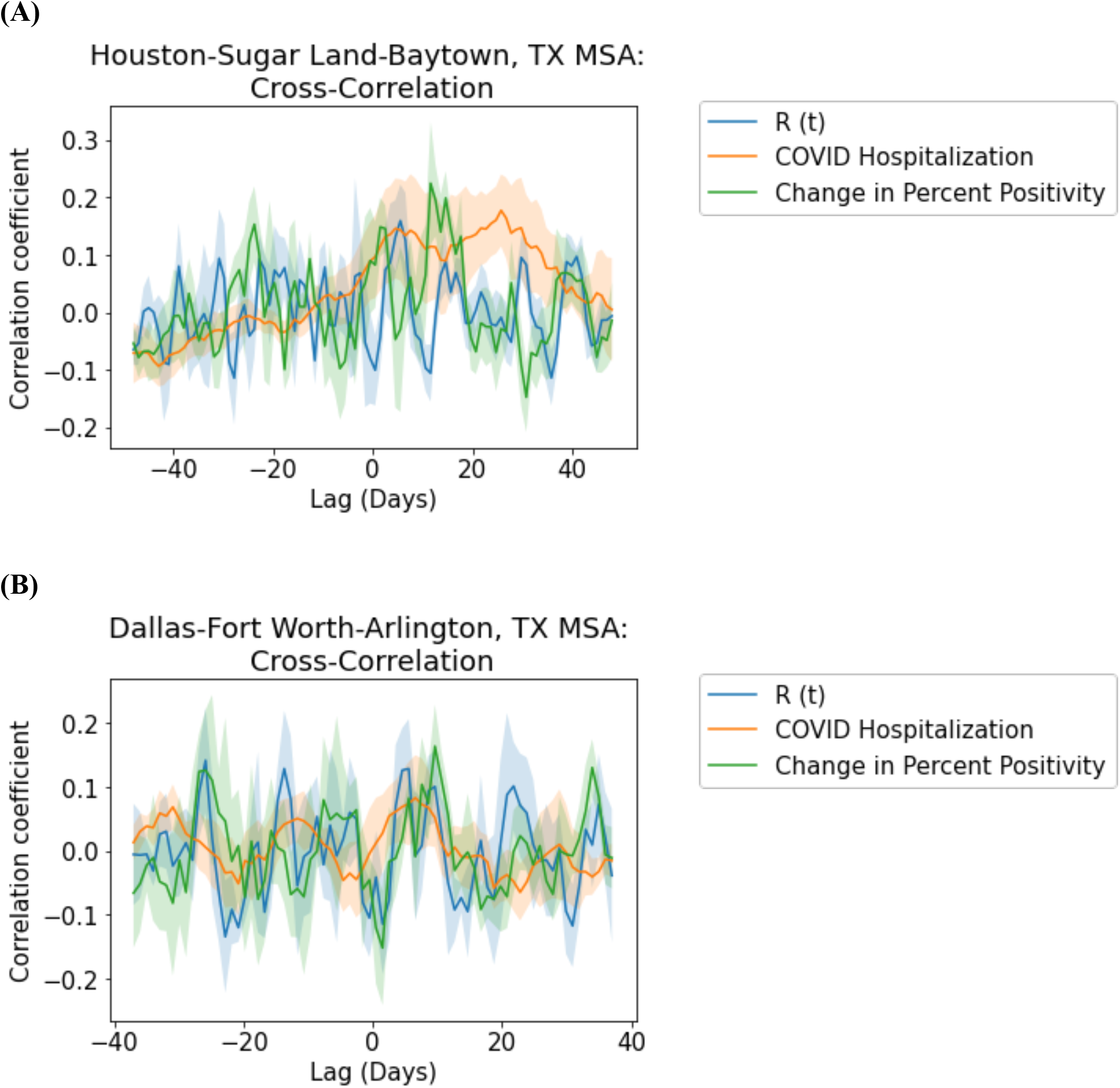
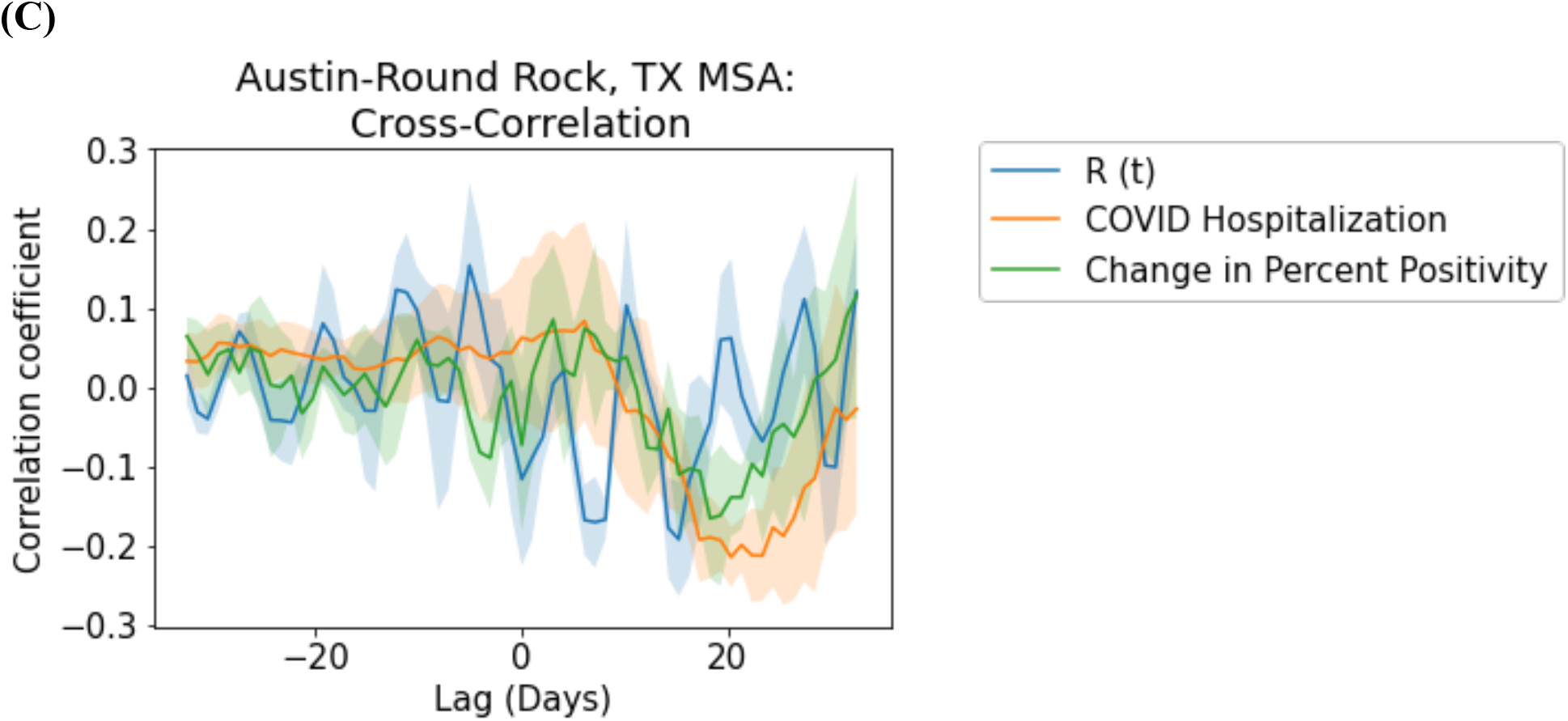
Cross-correlation plots of daily median cycle threshold (CT) values versus daily transmission rate R(t), daily number of individuals hospitalized with COVID-19, and daily change in percent positivity for (A) Houston-Sugarland-Baytown Metropolitan Statistical Area (MSA), (B) Dallas-Fort Worth-Arlington MSA, and (C) Austin-Round Rock MSA

**Supplementary Table 1.** Volume of tests per population in additional testing sites, compared to El Paso Metropolitan Statistical Area (MSA)

| Metropolitan statistical area (MSA) | Population | Number of tests conducted during study period | Tests per 100,000 individuals | Number of positive tests conducted during study period | Positive tests per 100,000 individuals |
| --- | --- | --- | --- | --- | --- |
| El Paso | 844,124 | 148,410 | 17,582 | 36,306 | 4,301 |
| Houston-Sugar Land-Baytown | 7,066,141 | 45,625 | 646 | 9,276 | 131 |
| Dallas-Fort Worth-Arlington | 7,573,136 | 34,279 | 453 | 9,539 | 126 |
| Austin-Round Rock | 2,227,083 | 16,730 | 751 | 1,177 | 53 |

**Supplementary Table 2.** Demographics for all samples sent to Dascena from each additional Metropolitan Statistical Area (MSA), compared to the El Paso MSA using a z-test with a significance level of p < 0.05. * p-value < 0.01 ** p-value < 0.001

|  | El Paso | Houston-Sugarland-Baytown | Dallas-Fort Worth-Arlington | Austin-Round Rock |
| --- | --- | --- | --- | --- |
| **Age (Mean, SD)** | 36.92 (18.53) | 37.48 (19.19) ** | 39.12 (19.23) ** | 35.78 (15.13) ** |
| **Gender** |  |  |  |  |
| Female | 81,520 (54.93%) | 25,369 (55.60%) | 19,564 (57.07%) ** | 9,022 (53.93%) |
| Male | 66,270 (44.65%) | 19,870 (43.55%) ** | 14,584 (42.54%)** | 7,396 (44.21%) NS |
| **Race** |  |  |  |  |
| White | 87,664 (59.07%) | 5,856 (12.84%) ** | 17,921 (52.28%) ** | 9,389 (56.12%) ** |
| American Indian or Alaska Native | 3,744 (2.52%) | 177 (0.39%) ** | 805 (2.35%) | 91 (0.54%) ** |
| Black/African American | 3,017 (2.03%) | 6,525 (14.30%) ** | 4,457 (13.00%) ** | 1,031 (6.16%) ** |
| Asian or Pacific Islander | 1,245 (0.84%) | 1,797 (3.94%) ** | 1,220 (3.56%) ** | 1,503 (8.98%) ** |
| Native American | 0 | 0 | 0 | 21 (0.12%) ** |
| Unknown | 52,203 (35.17%) | 30,599 (69.07%) ** | 9,604(28.02) ** | 3,742(22.37%) ** |
| **Ethnicity** |  |  |  |  |
| Hispanic | 127,722 (86.06%) | 20,648 (45.26%) ** | 17,198 (50.17%) ** | 4,907 (29.33%) ** |
| non-Hispanic | 8,632 (5.82%) | 21,250 (46.58%) ** | 13,991 (40.82%) ** | 8,820 (52.72%) ** |
| Unknown | 11,963 (8.06%) | 3,678 (8.06%) ** | 3,067 (8.95%) ** | 3,002 (17.94%) ** |

## REFERENCES

1. COVID-19 Dashboard by the Center for Systems Science and Engineering (CSSE) at Johns Hopkins University (JHU) [Internet]. COVID-19 Dashboard by the Center for Systems Science and Engineering (CSSE) at Johns Hopkins University (JHU). [cited 2021 Feb 16]. Available from: https://gisanddata.maps.arcgis.com/apps/opsdashboard/index.html#/bda7594740fd40299423467b48e9ecf6

2. US Food and Drug Administration. A Closer Look at COVID-19 Diagnostic Testing [Internet]. FDA. FDA; 2020 [cited 2021 Jan 27]. Available from: https://www.fda.gov/health-professionals/closer-look-covid-19-diagnostic-testing

3. RT-PCR Testing [Internet]. COVID-19 Real-Time Learning Network, by the Centers for Disease Control and Prevention and the Infectious Disease Society of America; 2020 [cited 2021 Jan 27]. Available from: https://www.idsociety.org/covid-19-real-time-learning-network/diagnostics/RT-pcr-testing/

4. Corman VM, Landt O, Kaiser M, Molenkamp R, Meijer A, Chu DK, et al. Detection of 2019 novel coronavirus (2019-nCoV) by real-time RT-PCR. Eurosurveillance [Internet]. 2020 Jan 23 [cited 2021 Jan 27];25(3). Available from: https://www.ncbi.nlm.nih.gov/pmc/articles/PMC6988269/

5. Tom MR, Mina MJ. To Interpret the SARS-CoV-2 Test, Consider the Cycle Threshold Value. Clin Infect Dis. 2020 Nov 19;71(16):2252–4.

6. Accelerated Emergency Use Authorization (EUA) Summary Modified Thermo Fisher TaqPath COVID-19 SARS-CoV-2 Test [Internet]. US Food and Drug Adminsitration; [cited 2021 Jan 27]. Available from: https://www.fda.gov/media/137450/download

7. Jefferson T, Spencer EA, Brassey J, Heneghan C. Viral cultures for COVID-19 infectious potential assessment - a systematic review. Clin Infect Dis Off Publ Infect Dis Soc Am. 2020 Dec 3;

8. Park JH, Jang JH, Lee K, Yoo SJ, Shin H. COVID-19 Outbreak and Presymptomatic Transmission in Pilgrim Travelers Who Returned to Korea from Israel. J Korean Med Sci. 2020 Dec 14;35(48):e424.

9. Salvatore PP, Dawson P, Wadhwa A, Rabold EM, Buono S, Dietrich EA, et al. Epidemiological Correlates of PCR Cycle Threshold Values in the Detection of SARS-CoV-2. Clin Infect Dis Off Publ Infect Dis Soc Am [Internet]. 2020 Sep 28 [cited 2021 Jan 6]; Available from: https://www.ncbi.nlm.nih.gov/pmc/articles/PMC7543310/

10. Jang S, Rhee J-Y, Wi YM, Jung BK. Viral kinetics of SARS-CoV-2 over the preclinical, clinical, and postclinical period. Int J Infect Dis. 2021 Jan 1;102:561–5.

11. Singanayagam A, Patel M, Charlett A, Bernal JL, Saliba V, Ellis J, et al. Duration of infectiousness and correlation with RT-PCR cycle threshold values in cases of COVID-19, England, January to May 2020. Eurosurveillance. 2020 Aug 13;25(32):2001483.

12. Bullard J, Dust K, Funk D, Strong JE, Alexander D, Garnett L, et al. Predicting Infectious Severe Acute Respiratory Syndrome Coronavirus 2 From Diagnostic Samples. Clin Infect Dis. 2020 Dec 17;71(10):2663–6.

13. Rao SN, Manissero D, Steele VR, Pareja J. A Systematic Review of the Clinical Utility of Cycle Threshold Values in the Context of COVID-19. Infect Dis Ther. 2020 Sep;9(3):573–86.

14. La Scola B, Le Bideau M, Andreani J, Hoang VT, Grimaldier C, Colson P, et al. Viral RNA load as determined by cell culture as a management tool for discharge of SARS-CoV-2 patients from infectious disease wards. Eur J Clin Microbiol Infect Dis. 2020 Apr 27;1–3.

15. Sarkar B, Sinha RN, Sarkar K. Initial Viral Load of a COVID-19-Infected Case Indicated by its Cycle Threshold Value of Polymerase Chain Reaction Could be used as a Predictor of its Transmissibility - An Experience from Gujarat, India. Indian J Community Med Off Publ Indian Assoc Prev Soc Med. 2020 Sep;45(3):278–82.

16. Jaafar R, Aherfi S, Wurtz N, Grimaldier C, Van Hoang T, Colson P, et al. Correlation Between 3790 Quantitative Polymerase Chain Reaction–Positives Samples and Positive Cell Cultures, Including 1941 Severe Acute Respiratory Syndrome Coronavirus 2 Isolates. Clin Infect Dis [Internet]. 2020 Sep 28 [cited 2021 Jan 6];(ciaa1491). Available from: https://doi.org/10.1093/cid/ciaa1491

17. Zhang X, Lu S, Li H, Wang Y, Lu Z, Liu Z, et al. Viral and Antibody Kinetics of COVID- 19 Patients with Different Disease Severities in Acute and Convalescent Phases: A 6-Month Follow-Up Study. Virol Sin. 2020 Dec 22;

18. Aslam A, Singh J, Robilotti E, Chow K, Bist T, Reidy-Lagunes D, et al. SARS CoV-2 Surveillance and Exposure in the Perioperative Setting with Universal testing and Personal Protective Equipment (PPE) Policies. Clin Infect Dis Off Publ Infect Dis Soc Am. 2020 Oct 22;

19. Liu Y, Yan L-M, Wan L, Xiang T-X, Le A, Liu J-M, et al. Viral dynamics in mild and severe cases of COVID-19. Lancet Infect Dis. 2020 Jun;20(6):656–7.

20. Choudhuri J, Carter J, Nelson R, Skalina K, Osterbur-Badhey M, Johnston A, et al. SARS-CoV-2 PCR cycle threshold at hospital admission associated with patient mortality. PloS One. 2020;15(12):e0244777.

21. Miranda RL, Guterres A, Lima CH de A, Filho PN, Gadelha MR. Misinterpretation of viral load in COVID-19. medRxiv. 2020 Oct 8;2020.10.06.20208009.

22. Binnicker MJ. Challenges and Controversies to Testing for COVID-19. J Clin Microbiol [Internet]. 2020 Oct 21 [cited 2021 Jan 26];58(11). Available from: https://jcm.asm.org/content/58/11/e01695-20

23. Hay JA, Kennedy-Shaffer L, Kanjilal S, Lipsitch M, Mina MJ. Estimating epidemiologic dynamics from single cross-sectional viral load distributions. medRxiv. 2020 Jan 1;2020.10.08.20204222.

24. Walker AS, Pritchard E, House T, Robotham JV, Birrell PJ, Bell I, et al. Viral load in community SARS-CoV-2 cases varies widely and temporally. medRxiv. 2020 Oct 27;2020.10.25.20219048.

25. Sheikhzadeh E, Eissa S, Ismail A, Zourob M. Diagnostic techniques for COVID-19 and new developments. Talanta. 2020 Dec 1;220:121392.

26. U.S. Department of Labor - Office of Workers’ Compensation Programs - Medical Fee Schedule [Internet]. [cited 2021 Feb 1]. Available from: https://www.dol.gov/agencies/owcp/regs/feeschedule/fee/fee11/READ_ME_FIRST_fs11_instructions

27. rtcovidlive [Internet]. GitHub. [cited 2021 Feb 1]. Available from: https://github.com/rtcovidlive

28. Bettencourt LMA, Ribeiro RM. Real time bayesian estimation of the epidemic potential of emerging infectious diseases. PloS One. 2008 May 14;3(5):e2185.

29. Texas COVID-19 Data [Internet]. Texas Department of State Health Services Texas COVID-19 Data. [cited 2021 Feb 4]. Available from: https://dshs.texas.gov/coronavirus/AdditionalData.aspx

30. Zein SE, El-Hor N, Chehab O, Alkassis S, Mishra T, Trivedi V, et al. Declining Trend in the Initial SARS-CoV-2 Viral Load During the Pandemic: Preliminary Observations from Detroit, Michigan. medRxiv. 2020 Jan 1;2020.11.16.20231597.

31. The Python Language Reference — Python 3.9.1 documentation [Internet]. The Python Software Foundation; 2021 [cited 2021 Jan 21]. Available from: https://docs.python.org/3/reference/

32. Mackey K, Ayers CK, Kondo KK, Saha S, Advani SM, Young S, et al. Racial and Ethnic Disparities in COVID-19–Related Infections, Hospitalizations, and Deaths. Ann Intern Med [Internet]. 2020 Dec 1 [cited 2021 Feb 11]; Available from: http://www.acpjournals.org/doi/full/10.7326/M20-6306

